# Delaying the COVID-19 epidemic in Australia: Evaluating the effectiveness of international travel bans

**DOI:** 10.1101/2020.03.22.20041244

**Authors:** Adeshina I. Adekunle, Michael Meehan, Dianna Rojas, James Trauer, Emma McBryde

## Abstract

Following the outbreak of novel Severe Acute Respiratory Syndrome Coronavirus-2 (SARS-CoV-2) or COVID-19 in Wuhan, China late 2019, different countries have put in place interventions such as travel ban, proper hygiene, and social distancing to slow the spread of this novel virus. We evaluated the effects of travel bans in the Australia context and projected the epidemic until May 2020. Our modelling results closely align with observed cases in Australia indicating the need for maintaining or improving on the control measures to slow down the virus.

## Introduction

As of 18^th^ March 2020, COVID-19 has caused almost 200,000 cases, 8,000 deaths and spread to over 150 countries (1). Declared a pandemic on 12^th^ March (2), it is clear that the world has lost the opportunity to contain the virus SARS-CoV2 as it managed to for SARS-CoV.

With 565 confirmed cases (3) and still counting, COVID-19 now looks certain to cause sustained local transmission within Australia. Therefore, at this time it is reasonable to reflect on the value of travel restrictions imposed to date, and to consider the benefit of ongoing travel restrictions in the coming weeks and months, when community transmission starts to increase. We answer these questions using OAG-travel data and a meta-population model for disease transmission. First we examine the counterfactuals: what would have happened had the travel ban from Wuhan/China not been implemented. Similarly we examine the impacts of bans to other emerging epicentres including Iran, Italy, and South Korea. We then examine the impacts for the future and compare the cases expected through community transmission, and from importation over the next 2 months.

## Methods

To model COVID-19 transmission we use a stochastic meta-population model, which categorizes the global population into susceptible, exposed, infectious or removed (SEIR) individuals. (See Appendix for full model specification). We parameterize the model to fit with observed case notification reports (4), allowing for un-notified cases. In China, we allow the reproduction number to be 2.63 from 1^st^ December, 2019 until 31^st^ January 2020, at which time extensive intervention measures successfully reduced the reproduction number to 1.73 (5). For future predictions, we consider both of these values to consider best- and worst-case scenarios which are applicable to Australia.

Migration patterns in our meta-population model are based on data obtained from OAG on international flight travel volumes for 200 countries in March 2018. From 1^st^ December 2019, until 24^th^ January 2020, we assume travel proceeds in accordance with these historic travel data. From 24^th^ January, we progressively impose travel restrictions, in accordance with IATA travel information.

We simulate our stochastic meta-population model 1000 times to generate estimates of: the cumulative number of imported cases in each country (see Figures 1 and 2) and the projected epidemic curves in Australia both in the presence and absence of travel bans.

**Figure 1.**
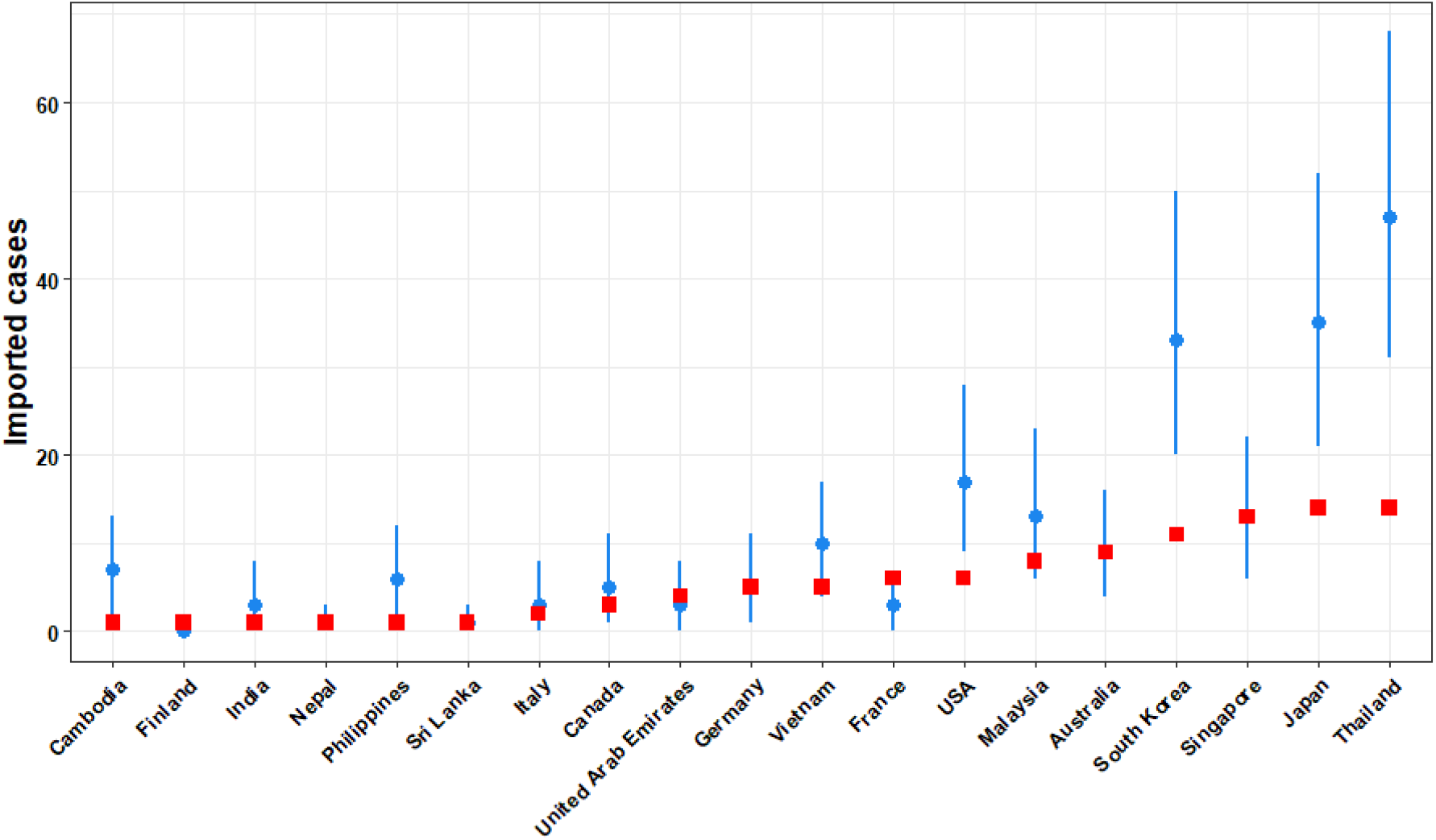
Cumulative imported cases of COVID-19 as at January 31, 2020 for the top 19 countries. The median cumulative importation and interquartile range are shown in blue, while the observed cases are shown in red.

**Figure 2.**
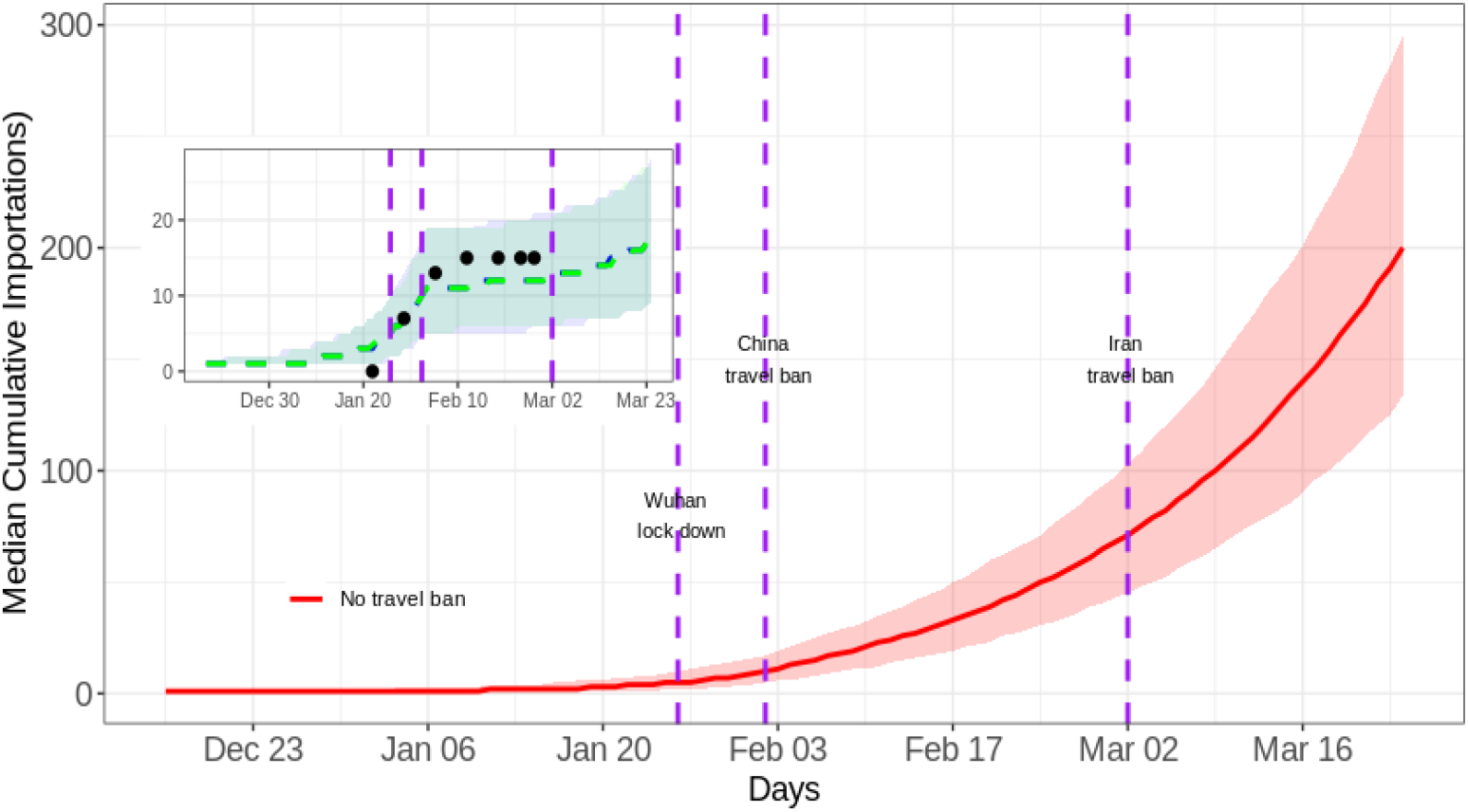
Travel Ban effect on COVID-19 importation to Australia. The red line shows the predicted cumulative number of importations up to 24^th^ March, 2020 if no travel bans had been enacted. (Inset) The reported (black dots) and predicted (green and blue curves) cumulative number of imported cases in the presence of travel bans (Travel ban on China, Iran, South-Korea and Italy). The blue dashed line shows the cumulative importations following both the Wuhan lock down and the travel ban placed on China. The green line is the effect of additional Iran, South Korea and Italy bans placed on the 29^th^ February and 5^th^ March, 2020 respectively.

## Results

We first validate our model by comparing estimates of imported cases provided by the model simulations with reported values as at January 31^st^ 2020 (see Figure 1). We find that our meta-population model accurately reproduces reported observations for many countries – in particular, Australia – however estimates become increasingly unreliable for countries with significant migration volumes with China. Variation between model predictions and observation can be partially accounted for by stochastic variation and potential under-reporting.

Figure 2 shows the counterfactual of the predicted number of cases Australia would have received if the ban on travel to China had not occurred. By 2^nd^ March 2020, our model estimates that Australia would have received over 70 imported cases of COVID-19 compared with the 15 cases that were actually observed. This represents a 79% reduction in expected cases, and similar to what was estimated elsewhere (6). However, the introduction of travel bans on international passengers arriving from Iran, South Korea and Italy, does not lead to a significant decrease in the expected COVID-19 importation count to Australia. One reason for this is that the much lower prevalence in these countries compared with China. Furthermore, Italy had already placed itself on lock down by the time Australia enforced restrictions on travelers arriving from Italy.

Finally, we explored the effects of travel ban and other intervention – such as reducing contacts to lower the basic reproduction number – on delaying widespread local transmission within Australia. We found that without travel bans, Australia would have experienced local transmission as early as Jan 15 and possibly have become the Pacific epicenter. However, with the China travel ban in place, this delayed the widespread occurrence of local transmission by approximately one month (see Figure 3). Thus, if interventions are in place that can reduce the reproduction number to 1.73 (in-line with China’s response) local transmission can be further delayed by another 5 weeks (Figure 3). Overlaying the simulation prediction with observed local cases in Australia suggesting more needs to be done to slow down the virus in Australia.

**Figure 3.**
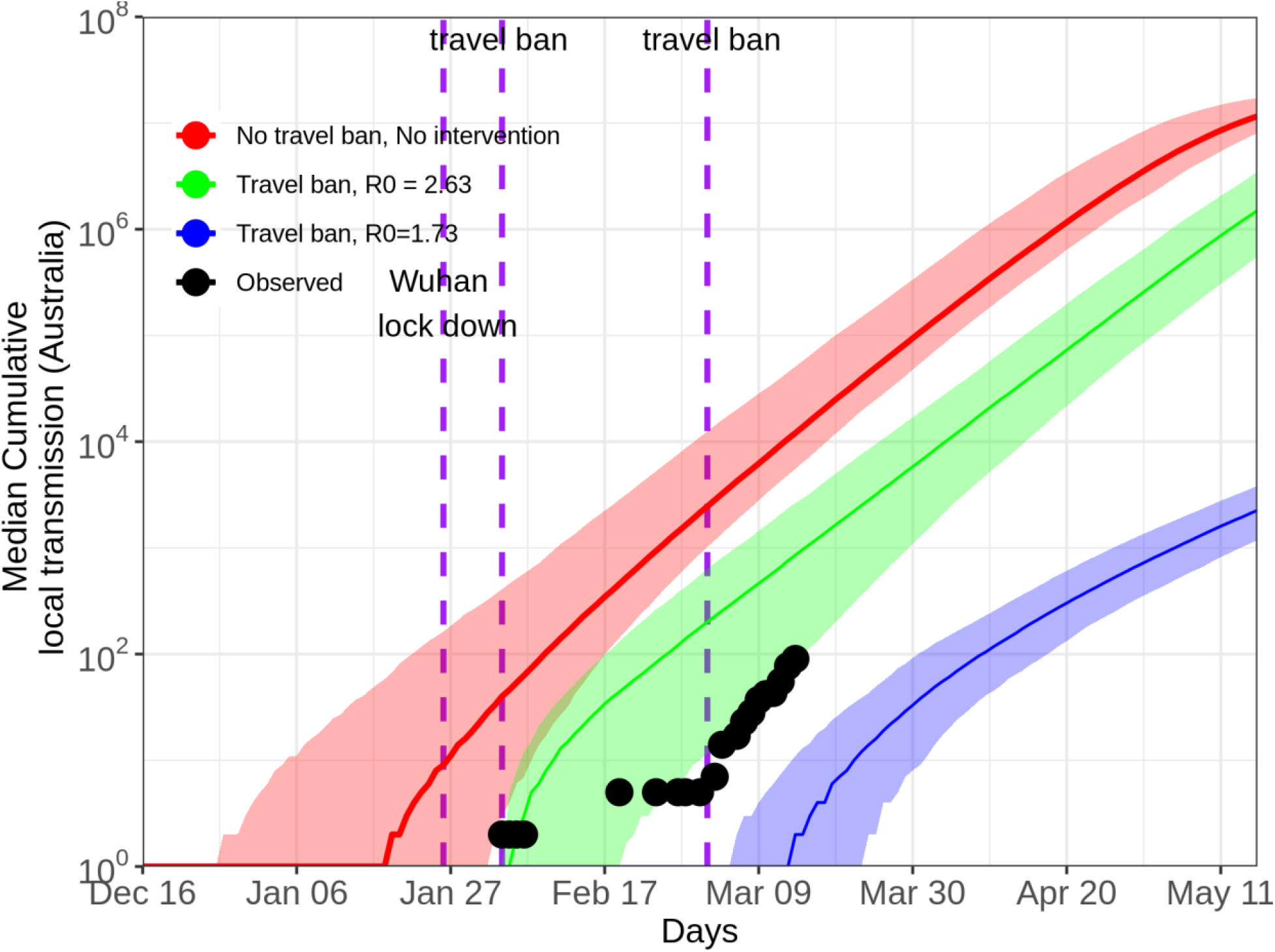
Effects of interventions on local transmission (Australia case study)

## Discussion

After an initial period of inactivity, many governments have now moved to impose international travel restrictions to prevent any further importation of COVID-19 cases. However, given that the virus is now a global pandemic and has reached most countries it is reasonable to question the impact of ongoing travel restrictions. In this short communication we have used international flight data and models of disease transmission to predict the national epidemic trajectory had travel bans not been put in place. Our results show that the travel ban on individuals arriving from China successfully delayed the onset of widespread transmission in Australia by four weeks. We also showed that travel bans from Hubei Province alone would have been much less effective, as the virus had already spread to many other provinces by the time the bans were enforced. Similarly, until now travel bans for South Korea and Iran (imposed on February 9 and March 5 respectively) were also shown to have negligible impact. However, as the number of cases in these countries continues to rise we expect these restrictions to become increasingly effective. Universal international travel bans coming into force now appear to be the only rational response, however, we need to consider how long they can and should last.

Future travel restrictions may have an impact for the next few weeks but potential importations will eventually be overwhelmed by local transmission, unless we can completely control transmission by achieving a reproduction number below 1. We estimate local transmission will outweigh imports sometime in the next two weeks if transmission proceeds at the pace it did in China prior to Jan 21 (R_0_ = 2.63), but will be delayed if Australia can successfully reduce transmission to a level similar to China from Jan 21 (R_0_ = 1.73).

## Data Availability

Data will be provided on request

## Appendix

**Fig. S2:**
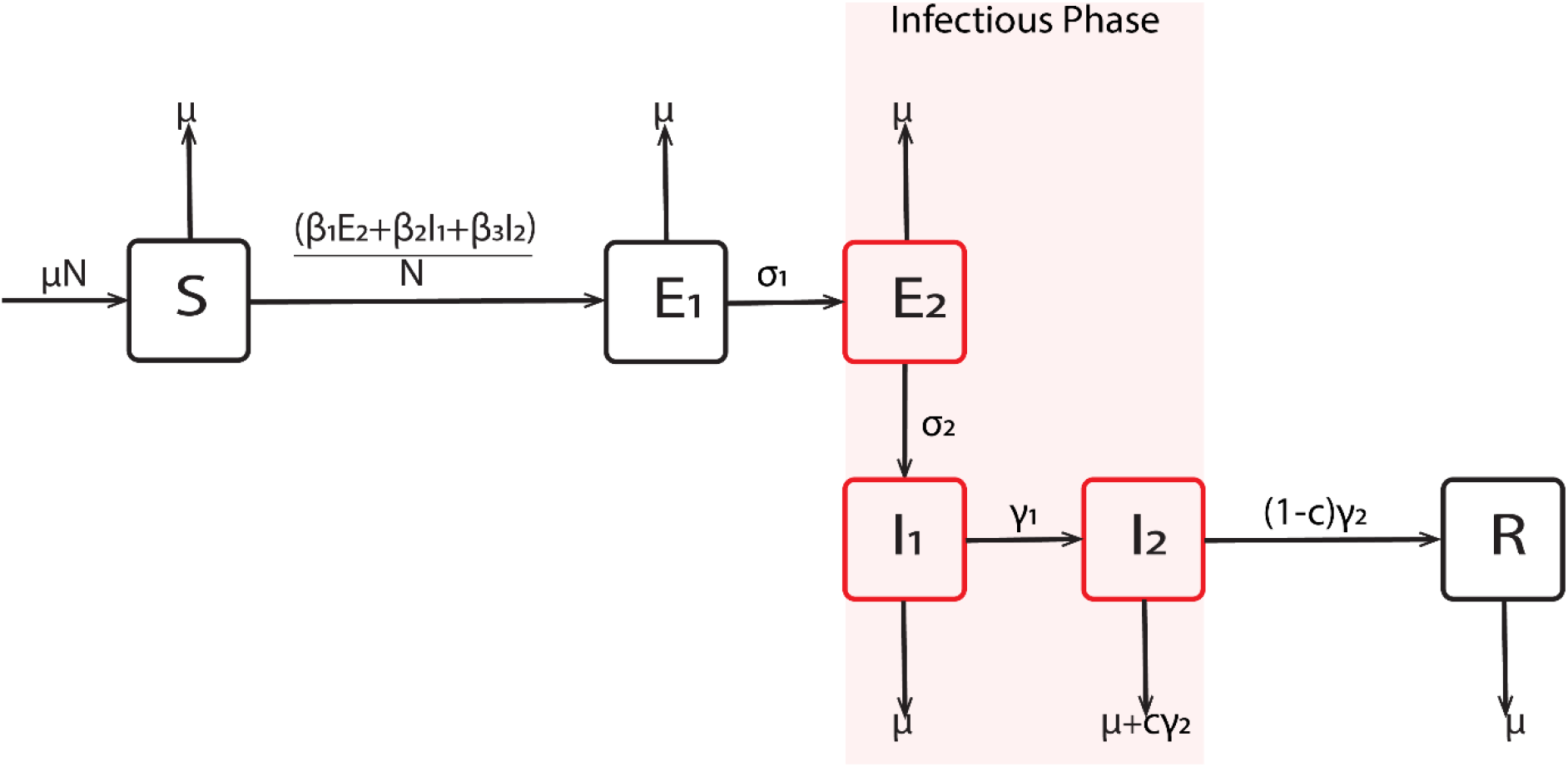
Schematic diagram of the transmission dynamics.

**Table S1:** State variable and parameter descriptions.

| Variable | Description |  |
| --- | --- | --- |
| $S$ | Susceptible class | |
| $E_1$ | Latent stage not infectious | |
| $E_2$ | Latent stage infectious | |
| $I_1$ | First stage of symptomatic | |
| $I_2$ | Second stage of symptomatic | |
| $R$ | Recovered class | |
| Parameter | Description | Values (references) |
| $\beta_1, \beta_2, \beta_3$ | Transmission rates for $E_2, I_1, I_2$ classes | 0.3125, 0.5, 0.176 ((7)) |
| $\sigma_1$ | First stage incubation rate | 0.3125 ((7)) |
| $\sigma_2$ | Second stage incubation rate | 0.5 ((7)) |
| $\gamma_1$ | First stage of recovery | 0.5 ((7)) |
| $\gamma_2$ | second stage of recovery | 0.176 ((7)) |
| $c_h$ | COVID-19 case fatality | 0.018 ((8)) |
| $\mu$ | Death rate | Not used |

Each country COVID-19 dynamics follows the schematic representation in Fig S2. The infectious classes are late latency *E*_2_ and the two stages of symptomatic infectiousness (*I*_1_ *and I*_2_). Patients either recover and moved to R class or die and are replaced to ensure constant population. The two-country dynamical model is as shown below:

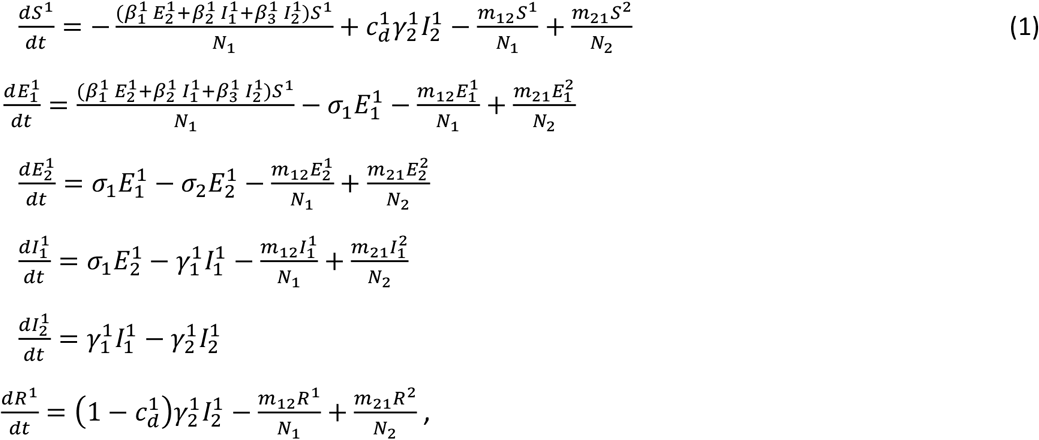

where superscript indicates country. This model is extended to all countries and coded in R (9) using the infectious disease node of the Australia Nectar Research cloud (www.nectar.org.au) as an individual-based model with a binomial distribution of the number of people that will experience an event at a specific time step. The events are infection, migration, emigration, recovery and death due to COVID-19. We neglected natural death, as this does not affect our result.

